# Experimental hybrid spectral CT with Cramér-Rao lower bound-optimized weighting for quantitative iodine imaging

**DOI:** 10.64898/2026.08.06.26359804

**Authors:** Olivia F. Sandvold, Roland Proksa, Amy E. Perkins, Heiner Daerr, Thomas Koehler, Tarun Jacob, Kevin M. Brown, Ewald Roessl, Peter B. Noël

## Abstract

Spectral computed tomography (CT) is a burgeoning quantitative imaging technique with applications in oncologic diagnostics, prognostic prediction, tissue perfusion studies, and treatment follow-up. While normalized iodine concentration values have been correlated with microenvironmental biophysical changes, obtaining accurate iodine concentrations, particularly at low concentrations remains difficult due to varying spectral CT instrumentation performance. Hybrid spectral CT systems, combining multiple spectral CT instrumentation techniques, address these quantitation insufficiencies by increasing spectral separation but have not been evaluated on a clinically analogous platform. We validate a hybrid spectral CT system, comprised of clinical-grade components, acquiring four distinct effective spectra and applying efficient noise-reducing weighting schemes to compare iodine noise and bias against conventional kVp-Switching (kVp-S). Two tube current levels (50, 350 mA) and three duty cycle ratios (33/67, 50/50, 75/25) were implemented to elucidate radiation dose exposure and kVp-S parameterization impact. A standard quality assurance (QA) and patient-derived, abdominal *IodinePrint* phantom were scanned on the system. The average absolute bias in iodine density images of the QA phantom was comparable across acquisition techniques, below 0.5 mg/mL, while quantitative noise improved by 22% using noise-optimized weighting schemes. In the *IodinePrint* phantom aorta and pancreas structures, the noise-optimized weighting scheme increased signal-to-noise ratio (SNR) by 1.3x compared to kVp-S alone. These results highlight the increased precision of hybrid, multi-channel spectral CT systems and motivate CT designs that enable robust CT biomarker development.

## 1. Introduction

Spectral computed tomography (CT) imaging not only has significantly improved image quality over conventional CT but also has bolstered the development of diagnostic iodine contrast workflows enabling earlier detection of tumors^1^, increased conspicuity of low concentration lesions^2^, and disease differentiation in oncology^3,4^. These advantages are enabled through the tissue-specific decomposition of two or more energy-dependent signals resulting in generation of iodine density^5–9^, virtual non-contrast (VNC)^10–12^, and virtual monoenergetic images (VMIs)^13–15^. While the benefits of acquiring spectral data have been demonstrated in various case studies^16,17^, current systems continue to be limited by both quantum noise (variance) and bias (error), which hinder accurate iodine quantification necessary for standardized biomarker development^18–20^. Although iodine density measurements have been correlated with histopathological features^21,22^, detecting subtle microenvironmental changes and identifying low-enhancing lesions remains a challenge^19,23,24^ and achieving uniformity across different spectral CT scanners depends critically on quantitative precision and accuracy^25,26^.

Currently, there are several technological approaches to spectral CT including dual-source^27,28^, rapid kVp-Switching (kVp-S)^29^, or slow spin-spin acquisitions, physical filters^30–32^ such as twin-beam filters, and detector-based hardware: dual- and triple-layer scintillators^33–35^ and direct-conversion photon-counting semiconductor materials^36–38^. Each technology has different spectral separation capabilities and inherent physical constraints that make unbiased, low noise signals difficult to obtain, particularly at low radiation doses and/or low iodine concentrations. To overcome the individual limitations of single instrumentation systems, it has been shown in prior experimental work that the combination of spectral technologies, enabling collection of three or more spectrally distinct datasets, can produce lower noise and more accurate results particularly for low concentrations of iodine^39–44^. To date, no experimental clinical grade “hybrid” system featuring source and detector hardware has been evaluated to validate these claims.

Despite their potential benefits, hybrid spectral CT systems which produce multiple spectral channels create increased processing demands to generate spectral images, limiting clinical feasibility. To address this issue, we implement previously presented novel weighting schemes to reduce multiple spectral channels into two material decomposition (MD) inputs to produce spectral results with optimal theoretical iodine noise^39^. The Cramér-Rao lower bound (CRLB) of variance in the iodine domain was implemented to predict system noise. These results demonstrated that the data reduction strategies estimated noise within 0.27% of an ideal, higher complexity four-channel MD approach. The noise-optimized channel reduction into two MD inputs is crucial for compatibility with most in-use clinical MD algorithms, which only require two spectral inputs to generate VMIs, VNCs, and iodine density images.

In this work, quantitative outcomes from an experimental hybrid spectral CT bench consisting of clinical-grade rapid kVp-Switching X-ray tube and dual-layer detector are presented. Specifically, comparisons of single instrumentation performance to data-reduced, multi-energy reconstruction techniques and elucidation of the impact of kVp-S parameters on quantitative metrics are evaluated. Two iodine-containing phantoms are used for bench evaluation—one standard quality assurance (QA) phantom and one anatomically realistic *IodinePrint* phantom capturing iodine-uptake in the pancreas and descending aorta.

## 2. Methods

### 2.1. Hybrid spectral CT bench components

To form the hybrid spectral CT bench system, a clinical-grade rapid kVp-S tungsten X-ray tube (vMRC, Philips Healthcare) and clinical-grade dual-layer detector (Spectral CT 7500, Philips Healthcare) were mounted with a source-to-detector distance of 1040 mm (**Figure 1A**). The collimation box (iCT, Philips Healthcare) contained two flat pre-patient filters: 2 mm aluminum and 1 mm titanium, and collimation was set to 80 mm at isocenter. The source-to-isocenter was 520 mm. A programmable stepper motor (PhyTron GmbH) was used to enable the rotation of 3D-printed patient-equivalent phantoms. The detector was equipped with a 2D anti-scatter grid. Finally, a fast photodiode was placed within the detector cradle in the path of the fan beam. The bench system recorded the tube voltage output, photodiode readings, and other system control states at 1 MHz.

**Figure 1.**
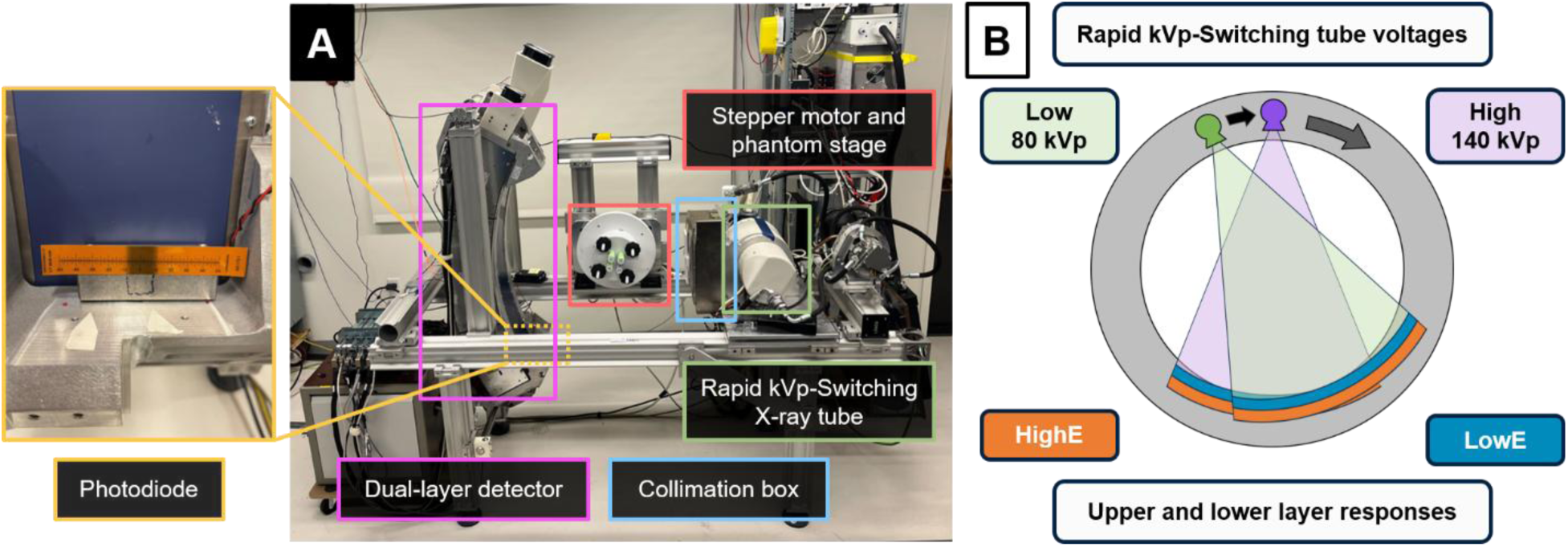
Photograph of bench system and schematic. (A) Hybrid, clinical-grade spectral CT system with photodiode detector and collimated X-ray beam shown on ruler. (B) Schematic of the system showing kVp-S high and low tube voltages with the corresponding dual-layer responses for each tube voltage.

The **Figure 1B** schematic demonstrates how two spectrally distinct, spatially aligned detector layers acquire simultaneous signals for each tube voltage level. For the energy-integrating dual-layer detector, the upper layer scintillator captured most of the low energy photons while the lower layer primarily measured high energy photons. The spectral channel data will be referred to with the following terminology:

**LowE**: Upper layer signal of the detector
**HighE**: Lower layer signal of the detector
**80 kVp**: Low kVp tube voltage
**140 kVp**: High kVp tube voltage

### 2.2. Image acquisition parameters

The rapid kVp-S X-ray tube was operated alternating between 80 kVp and 140 kVp tube voltages. Two tube currents of 50 and 350 mA were used to produce low and moderate radiation dose exposure levels. Given the cathode filament temperature cannot change as rapidly as the voltage, the generator maintained a stable temperature leading to greater X-ray fluence at high kVp compared to low kVp. The system automatically adjusted the filament temperature to produce the requested average mA. Cycle time (T_c_), the total duration of one pair of high and low voltage emission integration periods (IP), was set to 1000 microseconds [µs]. Three duty cycle ratios (DCs) of 33/67, 50/50, and 75/25 (140 kVp/80 kVp) were selected based on previous work^41,45^ to illuminate the impact of the total fraction of high kVp relative to low kVp projections on spectral noise and bias. The detector switched between high and low IP collection when a threshold tube voltage of 110 kV was passed during the transition periods.

Six kVp-S protocols were implemented using 50 and 350 mA tube currents: 33/67, 50/50, and 75/25 DC for each dose level. The actual achieved duty cycle ratio for each scan setting was measured using the duration of the kV waveform above and below the 110 kV threshold in 1000 cycle time periods centered around the temporal midpoint of the scan for each tube current and duty cycle pair. The average and standard deviation of high and low IP durations were computed over the sampled 1000 cycles. To approximate the rise and fall durations between high and low kVp, the voltage waveform for each of the sampled 1000 cycles was evaluated at the 110 kV threshold timepoints. The time difference between the first crossing of voltage thresholds at 135 and 85 kV determined the rise or fall time produced from the combination of tube current and duty cycle ratio.

### 2.3. Reconstruction and material decomposition

In each acquisition, four spectrally distinct projections or “spectral channels” were collected. They were: “80 kVp/LowE”, “80 kVp/HighE”, “140 kVp/LowE” and “140 kVp/HighE”. Pre-log detector intensities were collected from the phantom and air calibration scans, and three data reduction strategies were employed (**Figure 2**) to produce two inputs for material decomposition. These two inputs were used in 2D projection-based MD to generate spectral projections and subsequent spectral images.

**Figure 2.**
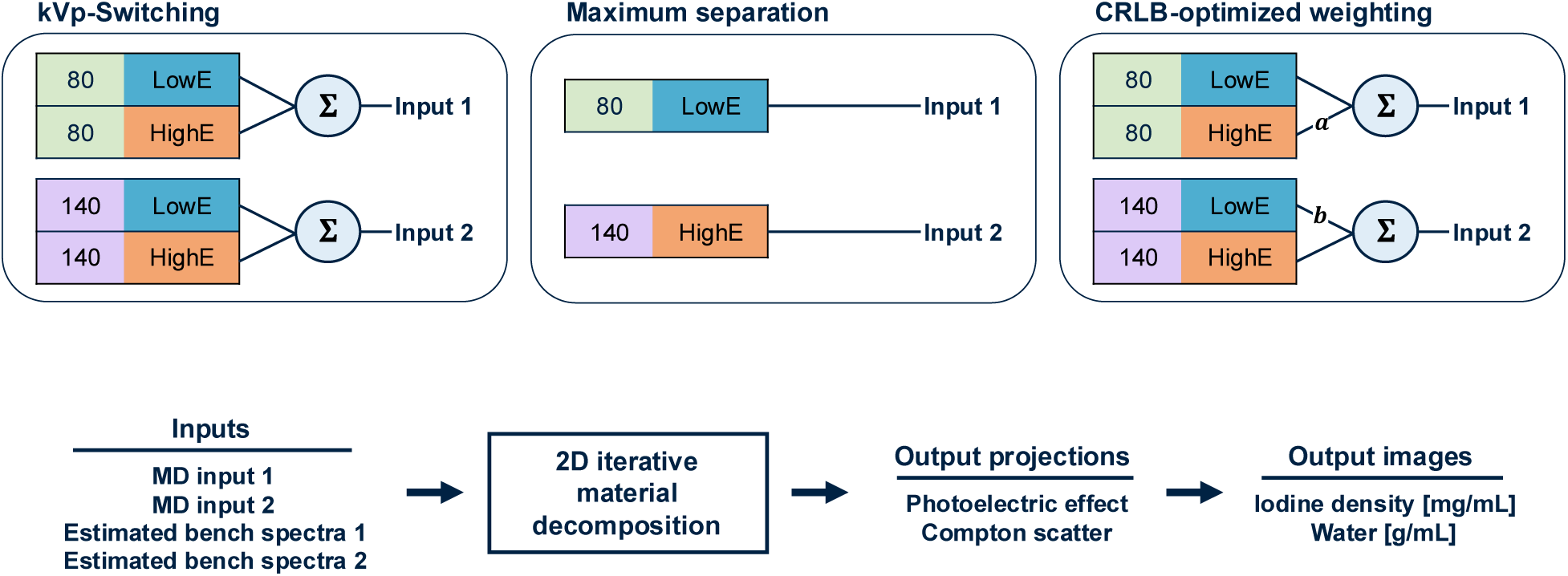
Reconstruction methods and MD process. (Top) Three approaches to generate two inputs for material decomposition. Weights ***a*** and ***b*** represent the contributions of the spectra and detector intensities into the MD inputs. (Bottom) Process to combine MD input projections with corresponding estimated bench spectra into iodine density and water basis images.

The first approach, **kVp-Switching** (kVp-S), used all four spectral channels by adding together the upper- and lower-layer intensities per kVp to mimic the behavior of a single-layer energy-integrating detector. The second approach, **maximum separation** (MaxSep), only used the two channels with maximum spectral separation from one another: 80 kVp/LowE and 140 kVp/HighE. In prior work, results demonstrated that the MaxSep method could produce similar CRLB iodine noise performance to an ideal four-input material decomposition^39^.

The last approach, a **CRLB-optimized weighting scheme**, selectively assigned weighted contributions of the four channels into two MD input projections by multiplying weights, *a* and *b*, that were computed to obtain the minimum estimated CRLB iodine for each scan protocol. The methodology to obtain these weights has been described fully in a previous publication^39^, but briefly described here: the spectral CT system was modeled in single-ray projection-space using switching characteristics of the X-ray tube including varying transition times and estimated current drop. Material bases CRLB variances were estimated after material decomposition. An iterative minimization search was performed to locate local weights, bound from [0,1], which produced the lowest CRLB iodine noise. The optimal weights for the six experimental protocols were determined using an input phantom path length of 200 mm water [1 g/mL] and 28 mm iodine vessel [10 mg/mL], closely matching QA quantitative experimental phantoms.

For all material decomposition schemes, the iodine CRLB signal-to-noise ratio (SNR) was estimated by dividing the simulated input iodine path length by iodine CRLB noise. MaxSep and CRLB-optimized scheme SNR values were compared to the kVp-S simulated CRLB iodine SNR.

Projection inputs 1 and 2 were formed after combining the spectral channel intensity data using the selected reconstruction method and estimated weights. An averaging filter was applied to the ten slices of collected sinogram data. The estimated spectral channel responses (Spectra 1 and 2) were generated using a system model of the X-ray tube, bench filtration, and detector sensitivities. By combining the direct sampling of the tube voltage and the indirectly estimated tube current derived from the photodiode signals, the effective spectrum for each input was calculated. These effective spectra included the switching voltage transitions contained in the cycle time duration. Correction for primary decay durations of a few microseconds in the upper and lower layer scintillator, respectively, were applied to the spectra to capture the spectral broadening effects of the primary decay arising from high kVp periods.

Using the expected model spectra and projection inputs, a 2D iterative projection-based material decomposition algorithm minimized least squares error and generated photoelectric effect (PE) and Compton scatter (CS) sinograms. Filtered back-projection (FBP) was applied to these spectral sinograms to generate PE/CS images which were linearly combined to produce iodine density and water maps^34^. The reconstruction field of view was 250 mm. No additional processing was implemented.

### 2.4. Standard spectral CT quantitative phantom

A polylactic acid (PLA) 20-cm diameter 3D-printed cylinder with an 80% infill ratio to match the Hounsfield unit (HU) of water was scanned (**Figure 3**). This phantom had slots for four Gammex (Sun Nuclear) tissue-mimicking inserts (Ø28 mm).

**Figure 3.**
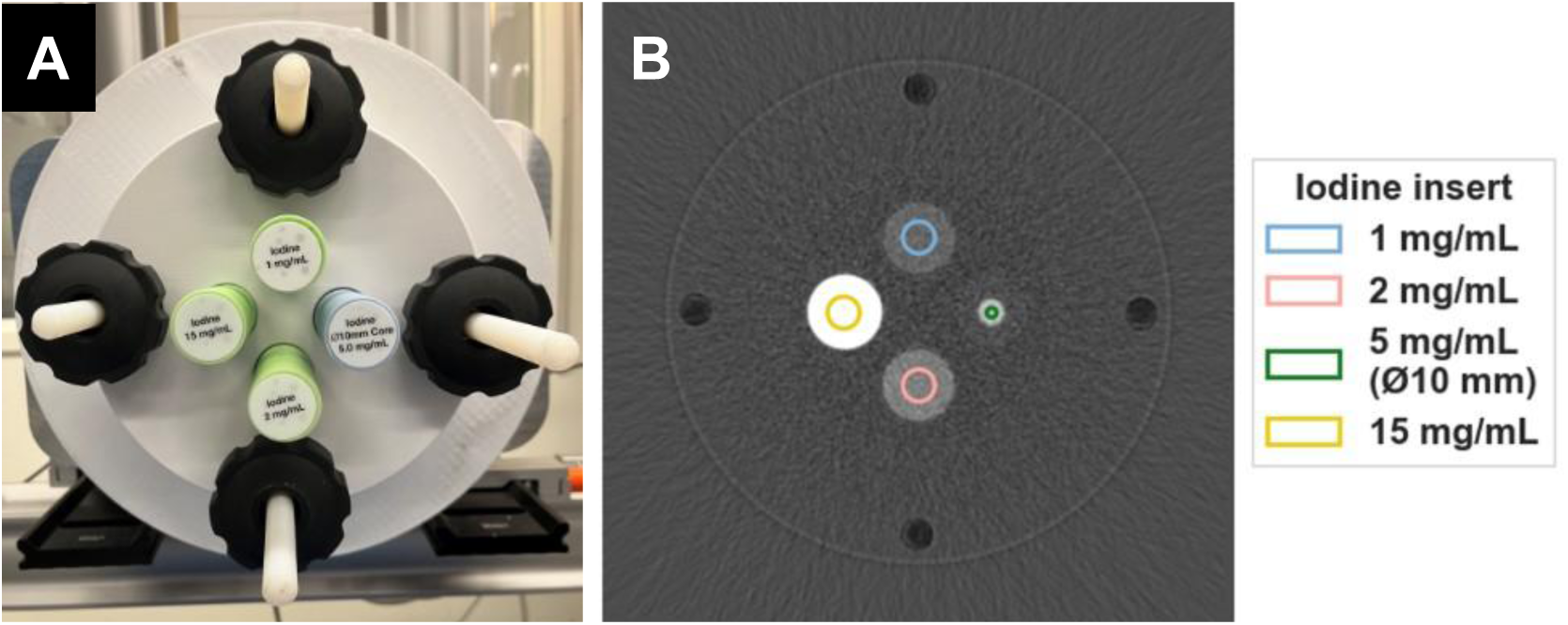
QA quantitative phantom. (A) 20-cm diameter PLA phantom containing tissue-mimicking inserts (configuration 1) mounted on rotation stage and (B) iodine density image with labeled ROIs. WW/WL: 10/2 mg/mL iodine.

To capture a range of physiological vessel contents and diameters, a total of seven inserts: iodine 1, 2, 5, 10, 15 mg/mL and solid water with a diameter of Ø28 mm and iodine 5 mg/mL with a diameter of Ø10 mm, were grouped into two configuration sets (**Table I**). The 1 mg/mL iodine insert was used in both sets for reproducibility.

**Table I.** Quantitative insert set configurations.

| Configuration 1 | Configuration 2 |
| --- | --- |
| Iodine 1 mg/mL | Solid water |
| Iodine 2 mg/mL | Iodine 1 mg/mL |
| Iodine 5 mg/mL (Ø10 mm) | Iodine 5 mg/mL |
| Iodine 15 mg/mL | Iodine 10 mg/mL |

For each protocol, one air scan and two phantom scans (one for each insert configuration) were collected.

The iodine density mean and standard deviation were measured on each image slice using circular regions of interest (ROI) placed in the center of each insert. The ROI diameter was 60% of the total insert diameter. Per slice iodine quantitative bias was calculated as the difference between measured concentration value and expected concentration calculated from the manufacturer’s composition specifications for each insert. The average noise and mean were computed over ten slices. The average absolute bias over iodine-containing rods was computed.

### 2.5. Patient-realistic, iodine-enhanced quantitative phantom

Further validation of the multi-energy reconstruction approaches was applied on a spectral, anatomically realistic pancreas phantom derived from a clinical CT patient acquisition. *PixelPrint* is a technology to translate patient attenuation images into reproducible, realistic 3D prints^46^. *IodinePrint* is a recent extension of *PixelPrint* to include multi-material filaments replicating contrast-enhanced studies. The fabrication process of the iodine-doped *IodinePrint* filament, dual-extrusion calibration, and comparison to expected values are outlined in a previous publication^47^. A 20-cm diameter *IodinePrint* phantom containing the liver, pancreas, descending aorta, and kidneys was scanned on the bench system at 1 Hz rotation, using 1000 µs cycle time, 33/67 duty cycle ratio, at 50 and 350 mA (**Figure 4**).

**Figure 4.**
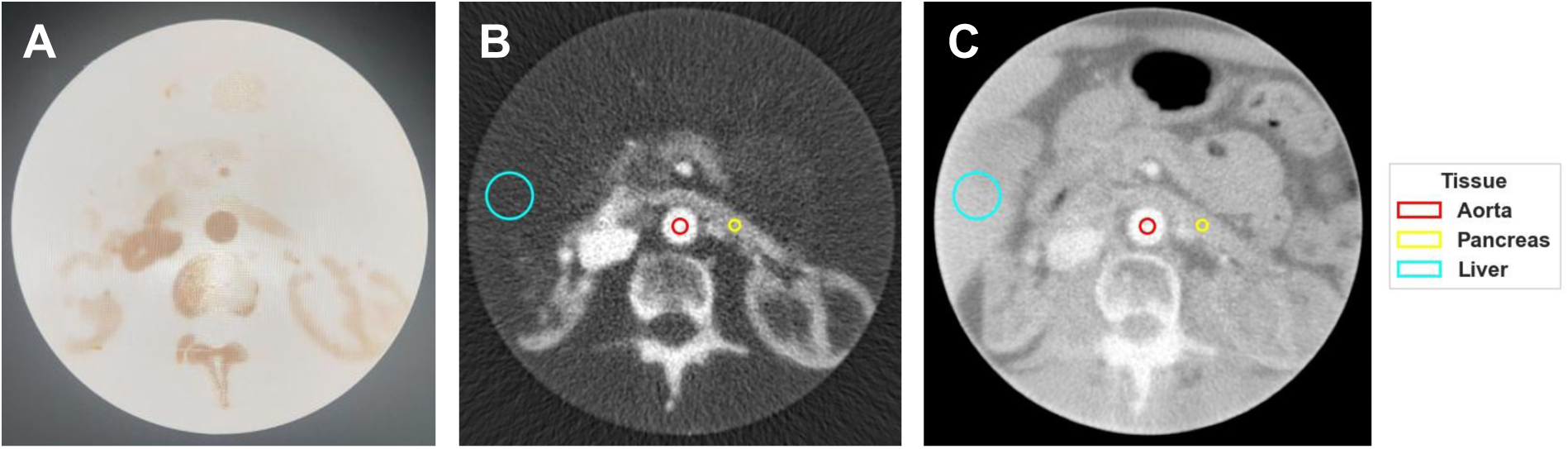
Quantitative, patient-realistic phantom. (A) Photograph of *IodinePrint* 20-cm diameter dual-filament, lifelike phantom. (B) Hybrid bench iodine density map with the aorta, pancreas, and liver background ROIs highlighted. WW/WL: 11/2 mg/mL iodine. (C) Water map from hybrid bench acquisition. WW/WL: 0.7/0.85 g/mL water.

The optimal weights for the CRLB-optimized scheme were generated with identical methodology in the previous section. For each reconstruction technique, the SNR was measured in one slice of the iodine density images by placing separate ROIs within the aorta and pancreas structures. The liver ROI was used for soft tissue background noise. A two-sided t-test was implemented to measure statistical significance (*p < 0.05*) comparing iodine SNR levels between reconstruction methods per mA per tissue type. From the initial patient iodine density image, the expected iodine concentrations within the structures was 12 mg/mL, 5 mg/mL, and roughly 1 mg/mL for the aorta, pancreas, and liver ROIs respectively.

## 3. Results

### 3.1. Optimal weights to minimize CRLB iodine noise

**Table II** showcases each reconstruction technique’s simulated iodine SNR and relative improvement of SNR compared to the kVp-S estimate using the same scan parameters. The highest estimated SNR is highlighted in green for each scan protocol. Across the two tube currents, increasing duty cycle ratio for 140 kVp led to decreasing kVp-S iodine SNR. In the higher dose 350 mA simulation, for 33/67 and 50/50 DC, the iodine SNR was ∼4x of the low tube current SNR regardless of MD method. In comparison, for the 75/25 DC, the 350 mA estimated SNR was around 3x greater than 50 mA estimates.

**Table II.** Optimal weights and estimated experimental iodine SNR.

| MD Approach | 50 mA, 33/67 DC |  |  | 350 mA, 33/67 DC |  |  |
| --- | --- | --- | --- | --- | --- | --- |
|  | Weights | Iodine CRLB SNR | Iodine SNR [rel. kVp-S] | Weights | Iodine CRLB SNR | Iodine SNR [rel. kVp-S] |
| kVp-S |  | 0.85 | 1.00 |  | 3.71 | 1.00 |
| MaxSep |  | 1.14 | 1.35 |  | 4.29 | 1.15 |
| CRLB-optimized | $a = 0.49$<br>$b = 0$ | 1.20 | 1.41 | $a = 0.59$<br>$b = 0$ | 4.51 | 1.21 |
| MD Approach | 50 mA, 50/50 DC |  |  | 350 mA, 50/50 DC |  |  |
|  | Weights | Iodine CRLB SNR | Iodine SNR [rel. kVp-S] | Weights | Iodine CRLB SNR | Iodine SNR [rel. kVp-S] |
| kVp-S |  | 0.83 | 1.00 |  | 3.41 | 1.00 |
| MaxSep |  | 1.12 | 1.35 |  | 4.00 | 1.17 |
| CRLB-optimized | $a = 0.49$<br>$b = 0$ | 1.18 | 1.42 | $a = 0.61$<br>$b = 0$ | 4.27 | 1.25 |
| MD Approach | 50 mA, 75/25 DC |  |  | 350 mA, 75/25 DC |  |  |
|  | Weights | Iodine CRLB SNR | Iodine SNR [rel. kVp-S] | Weights | Iodine CRLB SNR | Iodine SNR [rel. kVp-S] |
| kVp-S |  | 0.79 | 1.00 |  | 2.61 | 1.00 |
| MaxSep |  | 1.08 | 1.36 |  | 3.14 | 1.20 |
| CRLB-optimized | $a = 0.50$<br>$b = 0$ | 1.14 | 1.44 | $a = 0.62$<br>$b = 0$ | 3.39 | 1.30 |

All scan protocols showed the CRLB-optimized weighting scheme with the highest iodine SNR estimate, closely followed by MaxSep SNR performance. The optimal *b* weight for 140 kVp/LowE channel was 0 across all six scan protocols. Increasing tube current and increasing duty cycle ratio led to increased *a* weight values for the 80 kVp/HighE channel. The CRLB-optimized MD scheme was predicted to improve iodine SNR relative to kVp-S SNR by approximately 1.4x in 50 mA and by an average of 1.3x in 350 mA acquisitions.

### 3.2. Duty cycle vs tube current consistency

Figure 5 shows several switching voltage waveforms with 1000 µs T_c_ using various tube currents and duty cycle ratios. The average falling voltage duration for 50 mA was roughly 6x longer than the duration for the 350 mA acquisitions. In contrast, the rise time duration was consistently approximately 30 µs across tube current and DCs settings.

**Figure 5.**
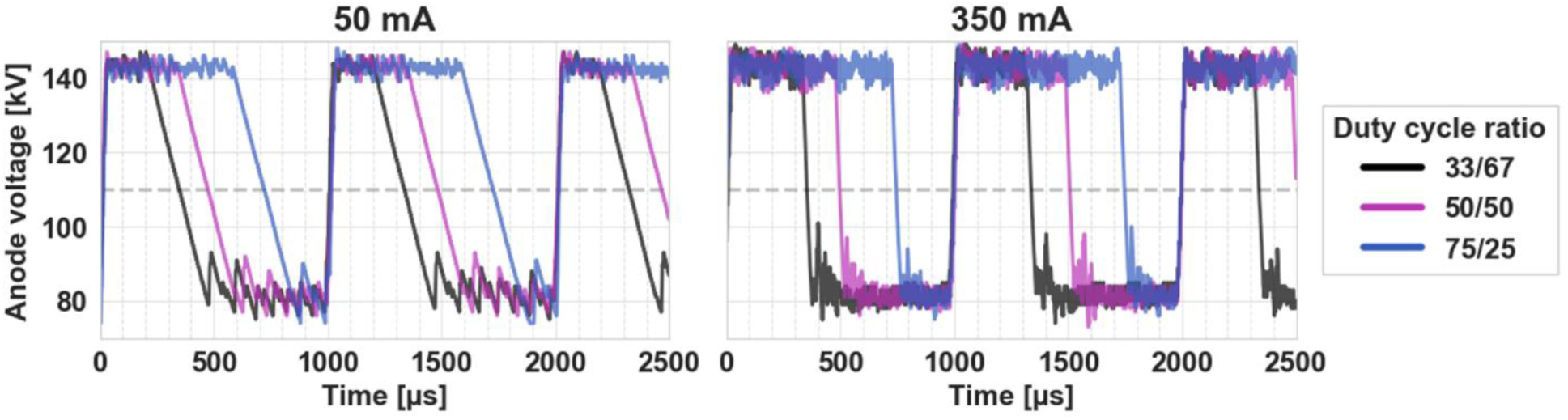
Tube voltage profiles. kVp-S tube voltage over time using varying tube currents and duty cycle ratios. The dashed gray line shows the IP transition threshold of 110 kV.

The average duration of T_c_ spent above and below 110 kV for each mA, duty cycle pairing is shown in **Table III**. The X-ray generator produced switching durations with less than 10 µs standard deviation over the 1000 sampled cycles.

**Table III.** High and low IP duration vs tube current and duty cycle setting.

| DC | 50 mA |  | 350 mA |  |
| --- | --- | --- | --- | --- |
| | High IP [μs] | Low IP [μs] | High IP [μs] | Low IP [μs] |
| **33/67** | $325 \pm 7$ | $674 \pm 7$ | $344 \pm 3$ | $655 \pm 3$ |
| **50/50** | $470 \pm 7$ | $529 \pm 7$ | $504 \pm 3$ | $495 \pm 3$ |
| **75/25** | $715 \pm 7$ | $284 \pm 7$ | $746 \pm 6$ | $253 \pm 6$ |

### 3.3. Quantitative iodine performance

The iodine density average noise in mg/mL, computed over all iodine containing inserts and all slices, versus the absolute average bias in iodine inserts per reconstruction method, per mA, and per DC is shown in Figure 6.

**Figure 6.**
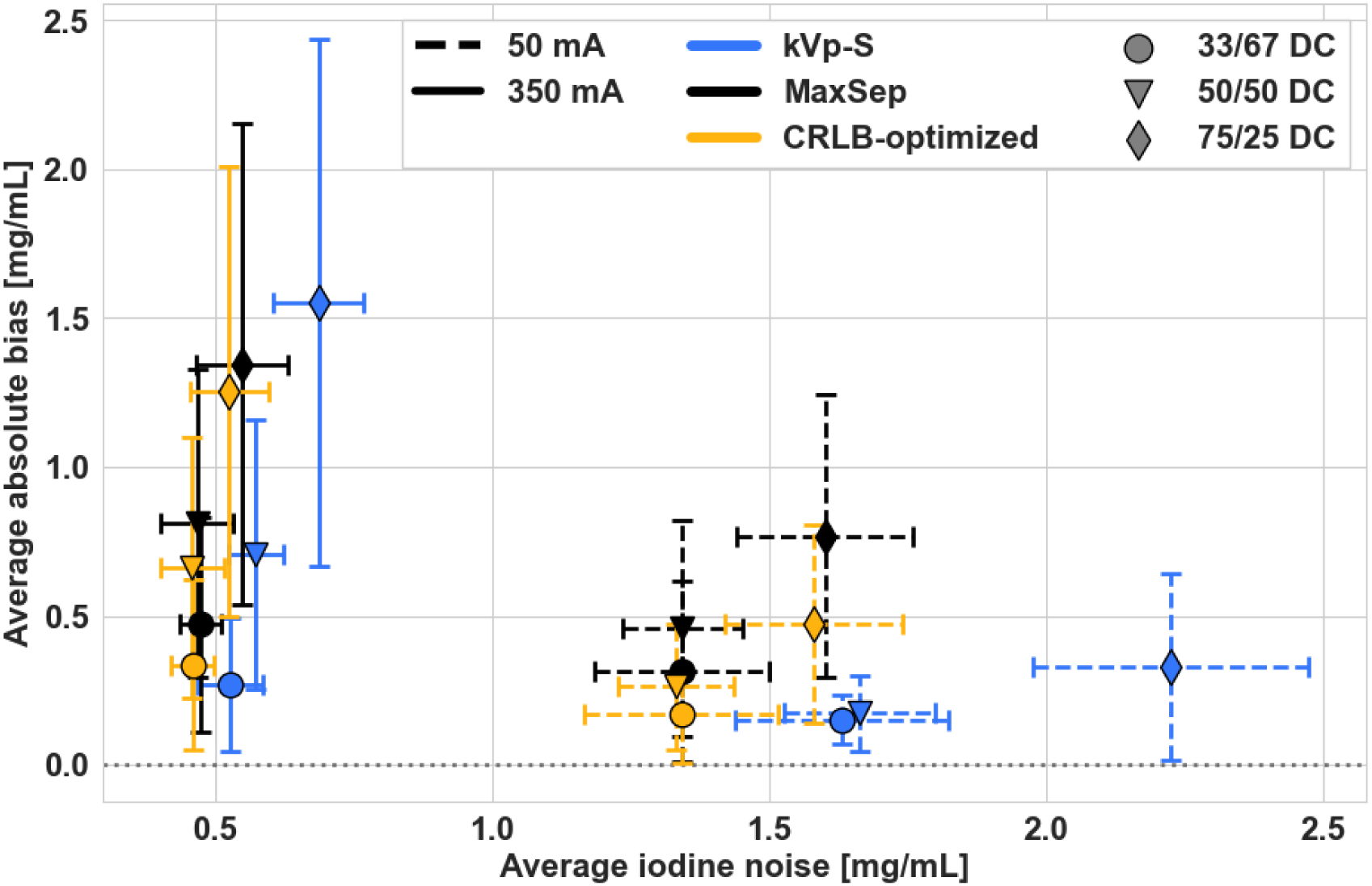
Quantitative measurements for multi-energy reconstructions. Mean measured iodine noise [mg/mL] versus average absolute bias [mg/mL] for 50 and 350 mA acquisitions at varying DCs and reconstruction techniques across slices and over all iodine containing inserts.

For 33/67, 50/50, and 75/25 DCs, the average iodine noise in the 50 mA acquisition iodine noise was greater than 350 mA noise by 3x, 3x, and 2x respectively. For both tube currents, increasing the DC increased noise for kVp-S reconstructions. In the 50 and 350 mA, 50/50 DC, CRLB-optimized recon, noise slightly decreased compared to the mA-matched 33/67 DC acquisitions. Within the tested DC/mA pairs, kVp-S reconstruction contained the highest average iodine noise of the MD methods. Changing the DC from 50/50 (triangle markers) to 75/25 (diamond markers) had a larger impact on iodine noise in 50 mA acquisitions compared to the difference in noise measured in the 350 mA scans as DC increased. Iodine noise in the CRLB-optimized MD was on average 22% and 19% less than kVp-S noise for 50 and 350 mA acquisitions across duty cycles while the average difference between MaxSep and kVp-S iodine noise was 22% and 16% less respectively.

The average absolute iodine bias over all iodine inserts was lowest in 33/67 duty cycle acquisitions for all MD methods. Using 350 mA, 33/67 DC, the absolute biases were 0.27 ± 0.2, 0.47 ± 0.4 and 0.34 ± 0.3 mg/mL for kVp-S, MaxSep, and CRLB-optimized MD schemes. The bias difference between kVp-S and MaxSep was statistically significant (*p < 0.001*) using a two-tailed pairwise t-test. In contrast, the difference between kVp-S and CRLB-optimized average absolute biases was not statistically significant (*p = 0.16*). In the 50 mA, 33/67 DC acquisition, the average absolute iodine errors were 0.15 ± 0.08, 0.32 ± 0.3, and 0.17 ± 0.2 mg/mL for kVp-S, MaxSep, and CRLB-optimized recons. Pairwise t-test analysis showed no statistically significant bias differences between kVp-S and the CRLB-optimized approach (*p = 0.51*). The 350 mA, kVp-S, 75/25 DC produced the largest iodine bias at 1.55 ± 0.9 mg/mL, but this error decreased by 13% and by 19% using MaxSep and CRLB-optimized schemes for the same acquisition. The 75/25 duty cycle also contained the largest bias variance, regardless of MD method.

### 3.4. Lifelike, quantitative *IodinePrint* phantom acquisition

Compared to kVp-S reconstructions, MaxSep and CRLB-optimized weighting scheme iodine density images contained increased SNR in both aorta and pancreas structures for 50 and 350 mA acquisitions (Figure 7).

**Figure 7.**
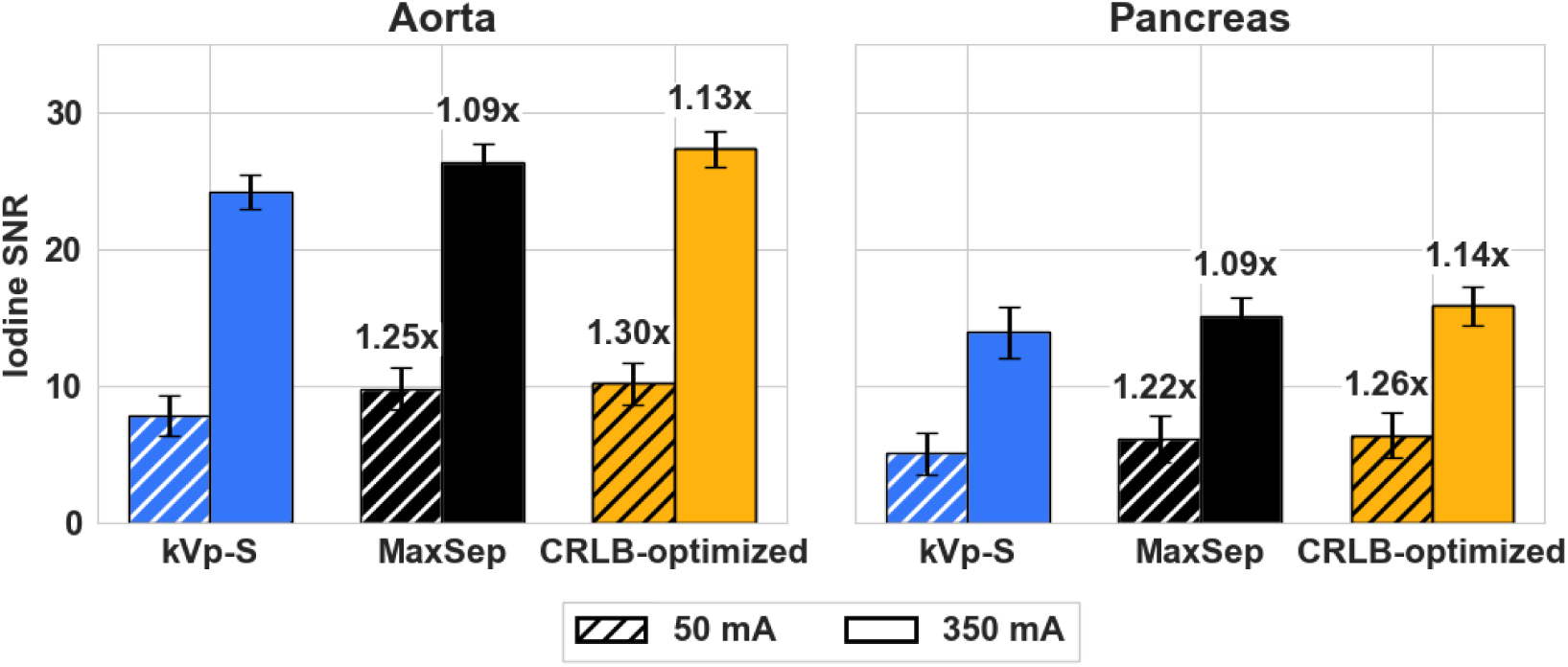
*IodinePrint* quantitative results. SNR in iodine density images for aorta and pancreas structures for two tube current levels and three reconstruction methods. The relative increases in SNR are labeled for MaxSep and CRLB-optimized against kVp-S acquisitions using the same tube current.

A two-tailed pairwise t-test between kVp-S/MaxSep and kVp-S/CRLB-optimized scheme found the differences in iodine SNR statistically significant (*p < 0.001)* for comparisons in aorta and pancreas tissue types and in all tube current settings. In 50 and 350 mA acquisitions, the CRLB-optimized iodine maps contained 1.3x and 1.1x SNR in tissue ROIs compared to kVp-S ROI SNR levels for those dose levels. Similarly, MaxSep recons yielded tissue SNR 1.2x and 1.1x the kVp-S SNR for the low and high tube currents. The 350 mA acquisition SNR, averaged over all the MD modes, was 2.8x and 2.6x the 50 mA scan SNR for aorta and pancreas ROIs respectively.

## 4. Discussion

We implemented an efficient data-reducing strategy to minimize iodine noise in spectral images on a clinical-grade experimental hybrid spectral CT bench at low and moderate dose levels using both a standard QA phantom and an anatomically realistic, iodine-containing phantom while modulating kVp-S acquisition parameters. We show that iodine noise significantly decreases when using a material decomposition CRLB-optimizing weighting scheme on multi-energy data while quantitative bias remains similar to kVp-S results. The measured noise improvements followed trends from single-ray projection-based CRLB simulations in both phantoms. In 50 and 350 mA scans, simulations predicted the CRLB-optimized scheme would reduce the noise by 28% and 19% respectively. Correspondingly, measured iodine noise values in the quantitative standard phantom were 22% and 19% less in CRLB-optimized scans compared to kVp-S at matched dose levels. Bench measurements of the *IodinePrint* phantom were 23% and 9% less, respectively. This increased quantitative precision obtained through hybrid spectral CT potentially supports the development of novel biomarker diagnostic workflows and large cohort correlation studies.

Observing the effect of duty cycle ratio, we found that favoring a balance between high and low kVp around 33/67 produced both low iodine bias (less than 10% error) and lower average iodine noise levels across a range of physiological iodine concentrations and vessel sizes. Comparing low and moderate dose levels, the iodine noise matched expectations to increase by a factor of roughly 2.6x in 50 mA images versus 350 mA acquisitions. The average iodine bias produced in kVp-S and CRLB-optimized schemes was similar within the paired duty cycle ratios.

For all scan protocols, the CRLB-optimized *a* weight was greater than 0.4 while the *b* weight was equal to 0. This indicates that the 140 kVp/LowE projection data should be excluded to produce minimal iodine noise in CRLB single-ray system models. As the duty cycle ratio increased, producing longer 140 kVp durations, the estimated *a* weight applied to the 80 kVp/HighE projection increased. This suggests that additional photons in MD input 1 are required to compensate for greater photons present in the 140 kVp/HighE MD input 2.

The impact of tube current magnitude on kVp-S transition times was clear in this study—a low tube current generated longer transition periods of up to 220 µs compared to fall durations of less than 40 µs in 350 mA scans. Increasing the tube voltage threshold (110 kV) that separates the high and low IP would result in greater duration of high energy X-ray beams in the low IP, affecting the detector SNR. Decreasing the difference in voltage potential pairs would decrease transition lengths at the cost of lower spectral separation. These settings (tube voltage threshold, kVp, tube current, DC, T_c_) not only influence the temporal and spatial resolution of images collected on the bench but also increase the optimization dimensions for such a hybrid system. While this study demonstrates the impact of 2D material decomposition inputs, the investigation of kVp-S parameters which enable fast temporal imaging or directly minimize bias in multi-material reconstructions are promising future directions.

Though the duty cycle ratio settings were implemented so that the 50 and 350 mA acquisitions would contain equal high and low IP durations, the average difference between measured IP durations was 28 µs across the three DCs, or 2.8% of the T_c_. Despite these differences, using the measured DCs, simulated CRLB-optimized weights did not change for either tube current setting. One limitation of this study is the exclusion of DCs with shorter 140 kVp IP durations, which have previously shown to contain less simulated expected material domain variance compared to two of the DCs in this study (50/50, 75/25)^48^. Additionally, only one pair of tube voltages (80/140 kVp) and one cycle time length (1000 µs) were investigated. To support potential clinical adoption of this technique, greater analysis of the hybrid system using T_c_ durations following clinical protocols and the combination of optimal DCs with both varying tube voltage pairs and radiation dose levels is necessary.

Several groups have demonstrated the benefits of a hybrid, multi-energy system. Simulation studies include work done by Tivnan et al. combining kVp-S, dual-layer detectors, and spatial-spectral (tiled array) filters into one CT system with improved iodine bias and noise compared to single instrumentations^44^, and by Wang et al. enhancing PCCT performance through the addition of a kVp-S source^40^. A cone-beam CT bench system was investigated by Zhou et al. using a fast kVp-S source stepped in 25 kV increments with a flat panel dual-layer detector^49^. Finally, there are examples of the application of optimized weights on multi-energy spectral datasets^50^ and data compression strategies on PCCT bin spectra^51^. This work fully integrates these various approaches on a flexible, clinical-grade spectral CT platform. We validated the simulated, hypothesized benefits of a hybrid, spectral CT system that combines optimizing weighting techniques and shows a significant improvement in iodine SNR compared to kVp-S alone. Furthermore, our bench can be equipped with filtration devices to increase spectral separation, and a milieu of lifelike, quantitative phantoms representing diverse clinical scenarios can be evaluated using the outlined experimental design.

In clinical practice, there are noise-bias tradeoffs which must be considered when designing optimization metrics. When lesion conspicuity and edge delineation is valued over quantitative accuracy, the application of weights to minimize CRLB material base noise is a practical approach. In contrast, when bias is a higher priority, the spectral channel weights could shift towards minimizing spectral estimation errors. There are several ways to estimate downstream quantitative bias including statistical methods^52^. Ultimately, methods such as the ones described in this paper provide more noise-efficient projection data which only serve to improve diagnostic spectral imaging combined with advanced reconstruction filtration, deep learning, and post-processing pipelines.

## 5. Conclusions

A clinical-grade, multi-instrumentation hybrid spectral CT bench was used to demonstrate efficient data-reduction material decomposition strategies to minimize iodine image noise in standard QA and lifelike spectral phantoms. As multi-energy spectral CT systems become more readily available, strategies to optimize image quality for task-specific diagnostic protocols while reducing system data burden are paramount for the clinical adoption of robust, accurate iodine biomarkers and high-quality spectral results including virtual non-contrast and iodine density images..

## Data Availability

All data produced in the present study are available upon reasonable request to the authors

## Acknowledgements

This work was supported through grants from the National Institutes of Health (R01EB030494) and Philips Healthcare.

## Notes

**Conflicts of interest:** AP, HD, TK, KB, and ER are employees of Philips. The authors have no other relevant interests to disclose.

### Competing Interest Statement

AP, HD, TK, KB, and ER are employees of Philips. The authors have no other relevant interests to disclose.

